# Supplementation with Berry Juice and Vitamin E Ameliorates Blood Cholesterol Level and Alters Gut Microbiota Composition

**DOI:** 10.1101/2023.05.22.23290321

**Authors:** Bangwei Chen, Yaxin Li, Zhiming Li, Xiaojie Hu, Hefu Zhen, Hongyun Chen, Chao Nie, Yong Hou, Xin Jin, Liang Xiao, Tao Li

## Abstract

**Scope:** Antioxidants, including vitamin E (VE) and grape seed extract, as anti-aging supplementation have been widely used to improve human health. However, the role of gut microbiota in dietary antioxidant supplementation is debatable. This study aimed to assess the longitudinal impact of dietary supplementation with antioxidant compounds on body health and the gut microbiota.

**Methods and results:** One hundred and twenty healthy individuals were randomly divided into a placebo group (amylodextrin) and three experimental groups ingesting different supplement (VE, grape seed extract, or mixed berry juice). Blood and fecal samples were collected during three intervention phases. We found that VE and mixed berry juice ameliorated blood cholesterol levels by reducing the levels of low-density lipoprotein cholesterol (LDL-C) in healthy volunteers. After the intervention, there was an increase in the relative abundance of short-chain fatty acid (SCFA)-producing bacteria and bile acid metabolizers. Specifically, the abundances of *Lachnospira* sp. and *Faecalibacterium* spp. increased in the VE and berry juice groups. Interestingly, the gut microbiota of poor responders harbored a greater proportion of disease-associated bacterial species.

**Conclusion:** Juice and VE could promote health by lowering LDL-C, partly and indirectly by affecting gut bacteria with the ability to produce SCFAs or metabolize bile acids.

## 1. Introduction

Population aging is accelerating, and several supplements, such as vitamin E (VE) and grape seed extract (GSE), have been generated and promoted as “anti-aging” products. In a systematic review including 884 randomized controlled intervention trials, it was suggested that supplementation with polyphenols may be beneficial for the heart; specifically, it was shown that supplementation with coenzyme Q10 could reduce overall mortality, while supplementation with β-carotene increased all-cause mortality. ^[1]^ However, the anti-aging mechanism of these products remains relatively unknown.

The efficacy of supplementation, the composition of which may include GSE, fish oil, VE, and/or vitamin C, is directly intertwined with the activity of the gut microbiota. ^[2, 3]^ It has been demonstrated that the relative abundance of bacteria within the genus *Roseburia*, which produces short-chain fatty acids (SCFAs), increases as a consequence of fermentation of VE in the human gut.^[4]^ In addition, studies have shown that the gut microbiota could degrade GSE into phenolic acids, which, in turn, promotes the relative abundance of SCFA-producing *Clostridium* spp. and induces an adaptive response to oxidative stress by activating the Nrf2 pathway.^[5-7]^ However, previous studies exploring the effect of anti-aging supplementation on gut microbiota composition have yielded contradictory conclusions.^[3]^

In the present study, we aimed to investigate the longitudinal effects of antioxidant supplementation on human health and their interplay with the gut microbiota. A three-month randomized controlled trial was adopted, and supplementation schemes involved the antioxidants VE, GSE, and mixed berry juice. None of the supplementation schemes affected relative telomere length, although VE and mixed berry juice ameliorated blood cholesterol levels by reducing the levels of low-density lipoprotein cholesterol (LDL-C) in healthy volunteers. These two supplementation schemes functionally altered the microbiome, and their influence was greatly weakened after a 3-month withdrawal period. In addition, the responses varied among the individuals, which could be partly explained by the baseline composition of the gut microbiota.

## 2. Experimental Section

### 2.1 Study design and sample collection

One hundred and twenty individuals were recruited on a voluntary basis to participate in this study based on the following eligibility criteria: i) aged 30 years or older; ii) not suffering from cancer, cardiovascular disease, or other chronic illnesses; and iii) not reporting antibiotic use within the last 30 days. The participants were randomly divided into four groups: i) juice (300 mL daily; juice composition: blueberry, 40%; apple, 40%; strawberry, 6%; cranberry, 7%; blackberry, 7%); ii) VE (400 IU/tablet, one tablet daily; GNC Holdings Inc., Pittsburgh, PA, USA); iii) GSE (300 mg/tablet, one tablet daily; GNC); or iv) placebo (corn amylodextrin, 300 mg/tablet, one tablet daily). Information on lifestyle and medical history was collected using a questionnaire. The exclusion criteria were discontinuity of supplement intake, antibiotic use, or inability to provide fecal samples for the study. Blood and fecal samples were collected at three different time points: at baseline (prior to the intervention), after the three-month intervention; and three months after a three-month withdrawal period. Blood and fecal samples were stored at −20 °C. The procedures involving human participants were reviewed and approved by the Institutional Review Board of BGI (No. BGI-IRB 20133). Affiliation: BGI-Shenzhen. All participants provided written informed consent to participate in the study.

### 2.2 Clinical laboratory testing and metabolic profiling

Blood samples were subjected to clinical laboratory tests at a licensed physical examination center. The tests included basic blood tests, such as the proportion of all cell types, and blood biochemistry tests. Targeted metabolomic profiling included amino acids, hormones, vitamins, microelements, and heavy metals. The methods for measuring the metabolites have been described previously.^[8]^

### 2.3 Metagenomic analysis of gut microbiota

DNA from fecal samples was isolated with the MGIEasy Kit from MGI and further processed with the MetaHIT protocol.^[9]^ Then, 500 ng of isolated DNA was used for library preparation and 100 bp single-read sequencing in the BGISEQ-500 platform.^[10]^ Low[_quality reads were removed using SOAPnuke version 1.5.2.^[11]^ Removal of human DNA sequences was conducted by subtracting reads that aligned against the GRCh38 human reference genome using Bowtie2 version 2.3.0.^[12]^ Alignment of mapped reads was performed using the Burrows-Wheeler Aligner (BWA) software version 0.7.17 against entries in the Unified Human Gastrointestinal Genome (UHGG) database (4644 strain-level complete genomes).^[13, 14]^ Contig depth of 4644 reference genomes was calculated using the script “jgi_summarize_bam_contig_depths” from Metabat version 2.12.1^[15]^ and normalized. Then, bacterial taxonomy annotation of assembled genomes was performed based on the Genome Taxonomy Database.^[16]^ Predicted functional analysis of genes from the reference genome was conducted using the Kyoto Encyclopedia for Genes and Genomes (KEGG)^[17]^ orthology (KO) database, and the relative abundance of each module was calculated based on KO abundance. Bacteria were considered to contain a module if at least half of the annotated KOs within a single module were mapped.

### 2.4 Statistical analysis

All statistical analyses were performed using R Statistical Software v4.1.0.^[18]^ The impact of potential confounding factors, such as age, sex, smoking, and drinking status, on gut microbiota features was evaluated using permutational multivariate analysis of variance (PERMANOVA) in R with the vegan package version 2.5-7^[19]^ based on the Bray-Curtis distance and 10,000 permutations.^[20]^ Species richness was calculated as within-sample diversity based on the Shannon index (alpha-diversity)^[21]^, whereas between-sample diversity was estimated based on Bray-Curtis dissimilarity^[22]^ and visualized by principal coordinates analysis (PCoA).^[23]^

Statistical tests, including the t-test, Wilcoxon rank-sum test, and nonparametric two-way ANOVA^[24, 25]^ were used to establish associations between the type of intervention, trial phase, and changes in the levels of blood metabolites or relative abundance of gut bacteria. False discovery rate (FDR) was used to control the rate of false positives. Nonparametric two-way ANOVA was performed using the ARTool package version 0.11.1 in R.^[26]^

Bayesian correlation analysis was conducted between bacterial strain logarithmic relative abundance and LDL-C levels considering the small sample size^[27]^ using the correlationBF function with 5000 iterations in the BayesFactor package version 0.9.12-4.4 in R.^[28]^

### 2.5 Data availability statement

The data that support the findings of this study are available from the CNGB Sequence Archive (CNSA)^[29]^ of the China National GeneBank Database (CNGBdb). Restrictions apply to the availability of these data, which were used under license for this study. Data are available from the corresponding authors with the permission of CNGBdb.

## 3. Results

### 3.1 Overview of randomized controlled interventional clinical trials

Out of 120 volunteers, 72 aged 32–69 completed the trial (average age of participants = 45.8 ± 10.0% years); specifically, 22 participants in the juice group, 12 in the VE group, 22 in the GSE group, and 16 in the placebo group (Figure 1A); 47.2% of participants were men (Table S1). Regarding fecal samples, 27 (37.5%) volunteers provided samples at the three sampling points, whereas 12 (16.7%) provided samples at the two sampling points. A total of 105 qualified samples were obtained and sequenced.

**Figure 1.**
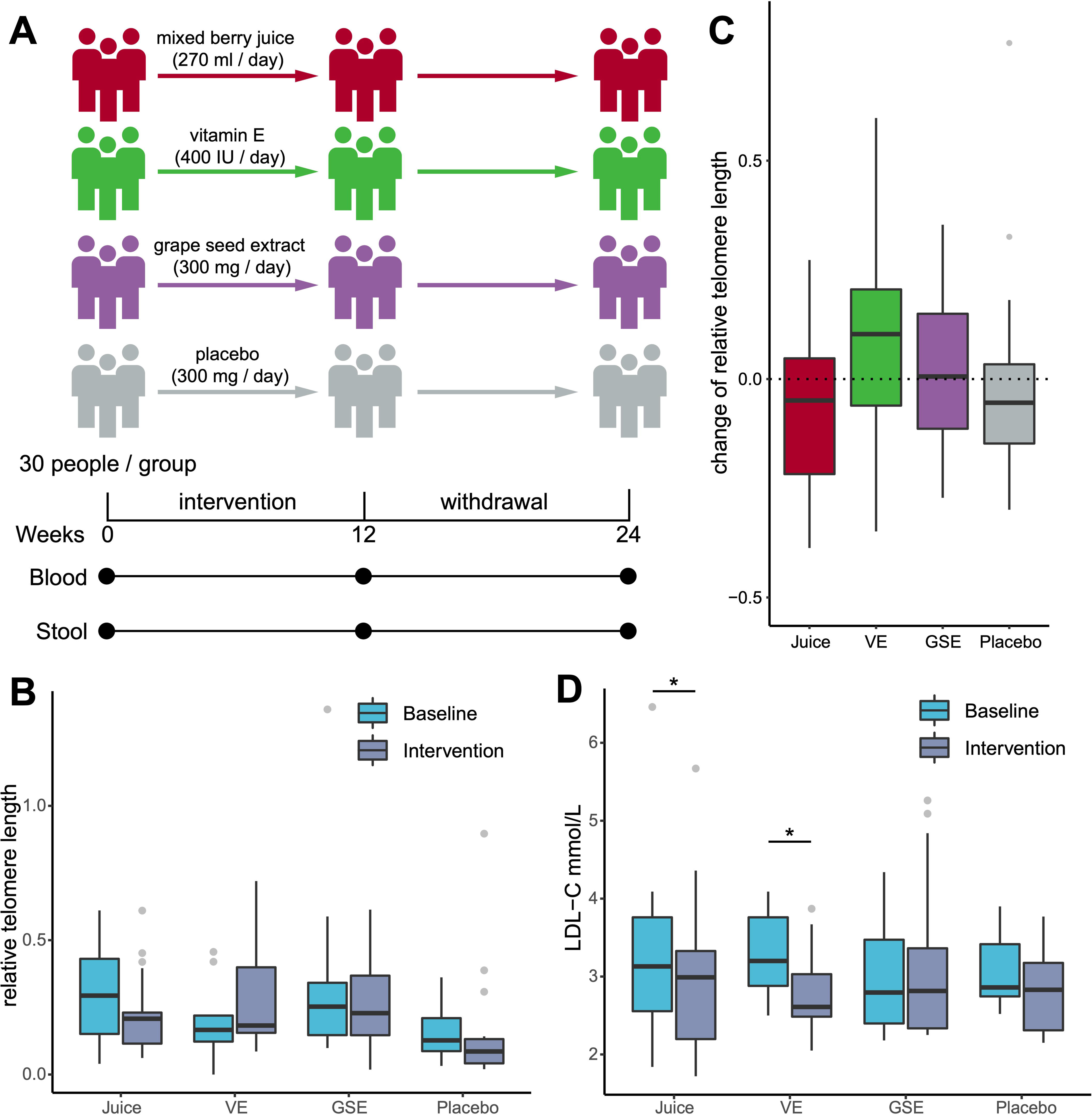
Study design and effect of supplementation schemes with antioxidants. (A) Schematic model of the study design. (B-D) Changes in relative telomere length and low-density lipoprotein cholesterol (LDL-C) levels during the intervention. Rank sum test and paired rank sum test were conducted to assess differences: (B, D) paired Wilcoxon rank-sum test and (C) Wilcoxon rank-sum test. \**P* < 0.05. VE, vitamin E; GSE, grape seed extract.

After the removal of human DNA and low-quality reads, an average of 1,192,247 high-quality reads were aligned to the reference microbial genomes, and an average of 88.9 ± 2.0% of reads in each sample were mapped (Table S2).

### 3.2 Juice and VE ameliorated lipid levels

To examine the effect of the supplementation schemes on aging, serum relative telomere length was measured as an aging biomarker.^[31]^ The relative telomere length did not change significantly before and after the intervention in any of the groups (*P*_paired Wilcoxon_ > 0.1; Figure 1B). Furthermore, changes in relative telomere length did not differ among the experimental groups (*P*_Wilcoxon_ > 0.1; Figure 1C). Immune and kidney ages were calculated as previously reported^[32]^, which did not change during intervention (*False discovery rate (FDR)*_paired t-test_ > 0.05), although GSE intervention decreased immune age (*FDR*_paired t-test_ = 0.096) and kidney age (*FDR*_paired t-test_ = 0.096).

Subsequently, 42 biochemical indicators were investigated in detail. In total, 19 indicators were altered in at least one group (*FDR*_Wilcoxon_ < 0.05; Table S3). None of the indicators in the placebo group changed (*FDR*_paired Wilcoxon_ > 0.1). In particular, inflammatory biomarkers, including absolute neutrophil count, neutrophil-to-lymphocyte ratio (NLR), and systemic immune-inflammation index (SII), were not significantly altered (*FDR*_paired Wilcoxon_ > 0.1). Considering the above changes (Figure S1), VE (*FDR*_paired Wilcoxon_ = 0.041) and juice (*FDR*_paired Wilcoxon_ = 0.015), but not GSE (*FDR*_paired Wilcoxon_ = 0.972), ameliorated lipid metabolism by reducing LDL-C levels (Figure 1D). LDL-C levels were the only indicator altered in the VE group. A previous study reported a significant decrease in serum triglyceride levels but not in LDL-C levels in an 8-week intervention with VE among patients with polycystic ovary syndrome.^[33]^

Moreover, the LDL-C levels were altered during the withdrawal period. LDL-C levels returned to levels similar to those found in the baseline group in both the juice group (*FDR*_paired Wilcoxon_ = 0.060; Figure S1A) and VE (*FDR*_paired Wilcoxon_ = 0.680) groups after the withdrawal period, but did not show a difference compared with the baseline level (*FDR*_paired Wilcoxon_ > 0.1). To determine whether the decrease in LDL-C levels depended on VE absorption, serum VE concentration was determined. As expected, the serum VE concentration increased after the intervention (*P*_paired Wilcoxon_ = 0.010) and decreased after the withdrawal period (*P*_paired Wilcoxon_ = 0.002; Figure S2A). Nonetheless, no obvious correlation was observed between the rate of change in VE and LDL-C (*P*_Spearman_ = 0.634; Figure S2B). Subsequently, the relationship between baseline status and the rate of change was evaluated, which revealed that the absolute rate of LDL-C change may be positively correlated with the baseline serum VE level (*rho* = 0.554; *P*_Spearman_ = 0.082).

### 3.3 Microbial changes associated with supplementation schemes impacted LDL-C levels

Given the decrease in LDL-C levels in participants after supplementation with VE or mixed berry juice, it was speculated whether dietary supplementation resulted in a persistent change in participants’gut microbiota and whether these alterations strengthened the hypolipidemic effect of the supplemented compounds. Therefore, strain-level annotation was performed using the UHGG database. In total, 841 bacterial strains were detected in 19 volunteers (11 in the juice group and 8 in the VE group). Considering that host factors may affect the gut microbiota, PERMANOVA was used, and the results showed that age (*P* _PERMANOVA_ = 0.309) and sex (*P* _PERMANOVA_ = 0.147) did not affect the composition of the baseline gut microbiota in healthy individuals.

Considering the gut microbiota structure, strain-level alpha diversity (Shannon diversity index) and richness did not differ throughout the intervention phases in the VE and berry juice groups (*P*_paired Wilcoxon_ > 0.05; Figure S3A, S3B; Table S4). For reliability, we further analyzed the composition of the gut microbiota at the genus and species levels, and found that Shannon diversity and richness were not significantly altered after the intervention (Table S4). In addition, the beta diversity of gut bacterial communities was evaluated using a PCoA plot with Bray-Curtis dissimilarity (Figure S3C), and Bray-Curtis distances among samples were compared. Although the distance between samples within the same group did not change as a result of supplementation (*P*_Wilcoxon_ > 0.05; Figure S3D; Table S5), a lower distance was found between samples in the VE group compared to those in the juice group (*P*_Wilcoxon_ < 0.001). Additionally, trajectory analysis based on data related to the abundance of bacterial strains was performed, and the results showed that the microbiota of individuals returned to baseline status during the withdrawal period, as expected (Figure S4, S5).

Subsequently, we investigated whether supplementation led to the enrichment of certain bacterial strains and whether these could be related to LDL-C levels. The number of fecal samples evaluated in this study was limited; thus, the significance threshold alpha was set to 0.1. Comparing the relative abundance of 841 bacterial strains, 40 bacterial strains were found to be significantly altered in the juice group (*P*_paired Wilcoxon_ < 0.1; Table 1; Figure 2A), whereas 10 bacterial strains were found to be significantly altered in the VE group (*P*_paired Wilcoxon_ < 0.1; Table 2; Figure 2B). We further applied Bayesian correlation instead of classical Spearman correlation to mine LDL-C-related bacterial strains, and two additional differential bacterial strains were found, namely *Monoglobus pectinilyticus* (juice group; |*r|* > 0.34; *P*_Bayesian correlation_ < 0.1, Figure S6A, S6B) and UBA9502 sp003506385 (VE group; |*r|* > 0.39; *P*_Bayesian correlation_ < 0.1, Figure S6C, S6D), which were negatively correlated with LDL-C levels at the baseline and intervention stages. Supplementation with juice and VE led to an enrichment of *Lachnospira* sp. and *Faecalibacterium* spp. in line with previous reports, which showed that these two bacterial species were depleted in obese subjects and were negatively associated with LDL-C levels.^[34-37]^

**Figure 2.**
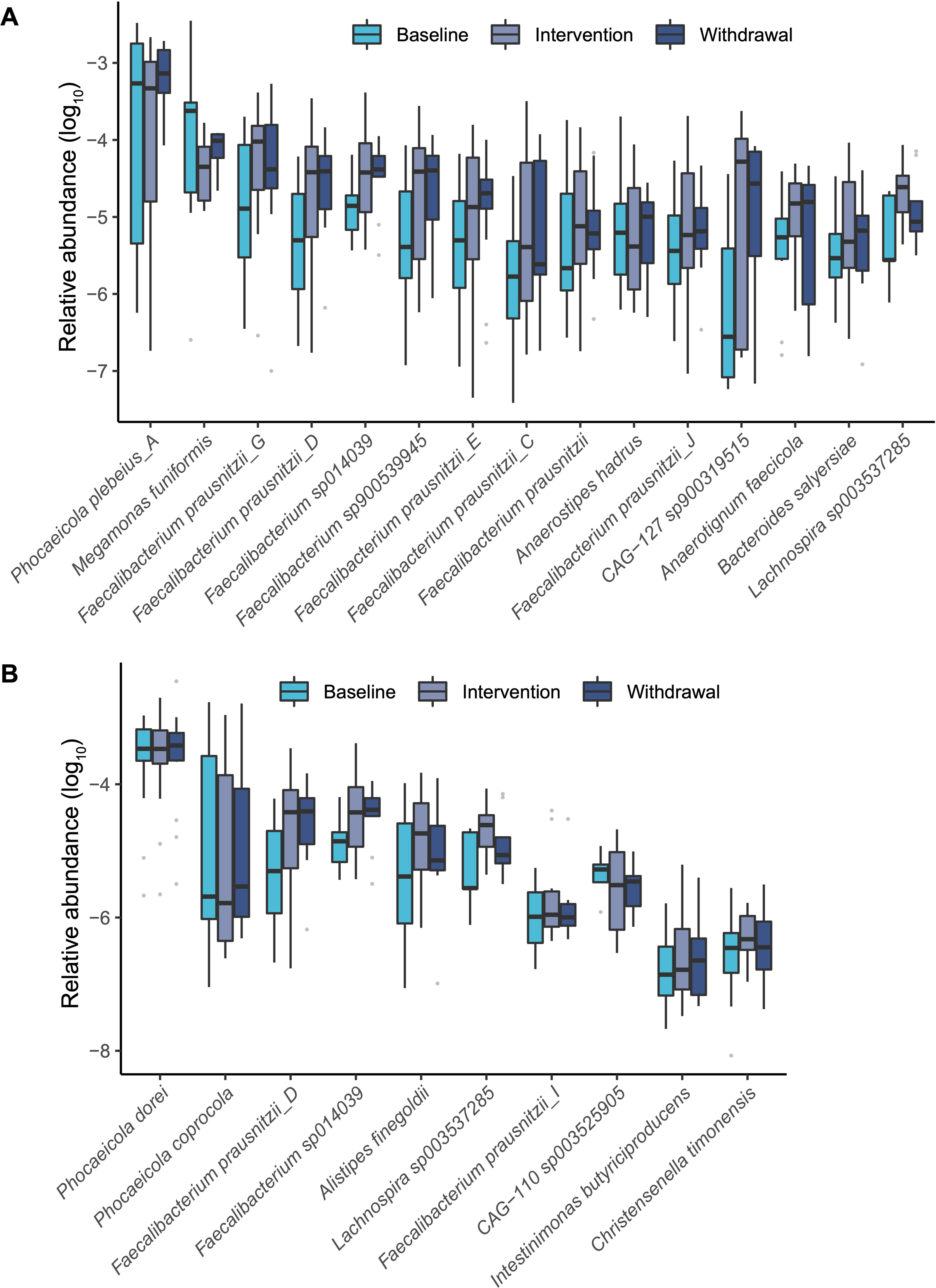
Compositional changes in the gut microbiota of healthy individuals receiving supplementation with antioxidants. (A) Log_10_ of the relative abundance of the top 15 bacterial strains in individuals in the mixed berry juice group. (B) Log_10_ of the relative abundance of the bacterial strains in individuals in the vitamin E group.

**Table 1.**
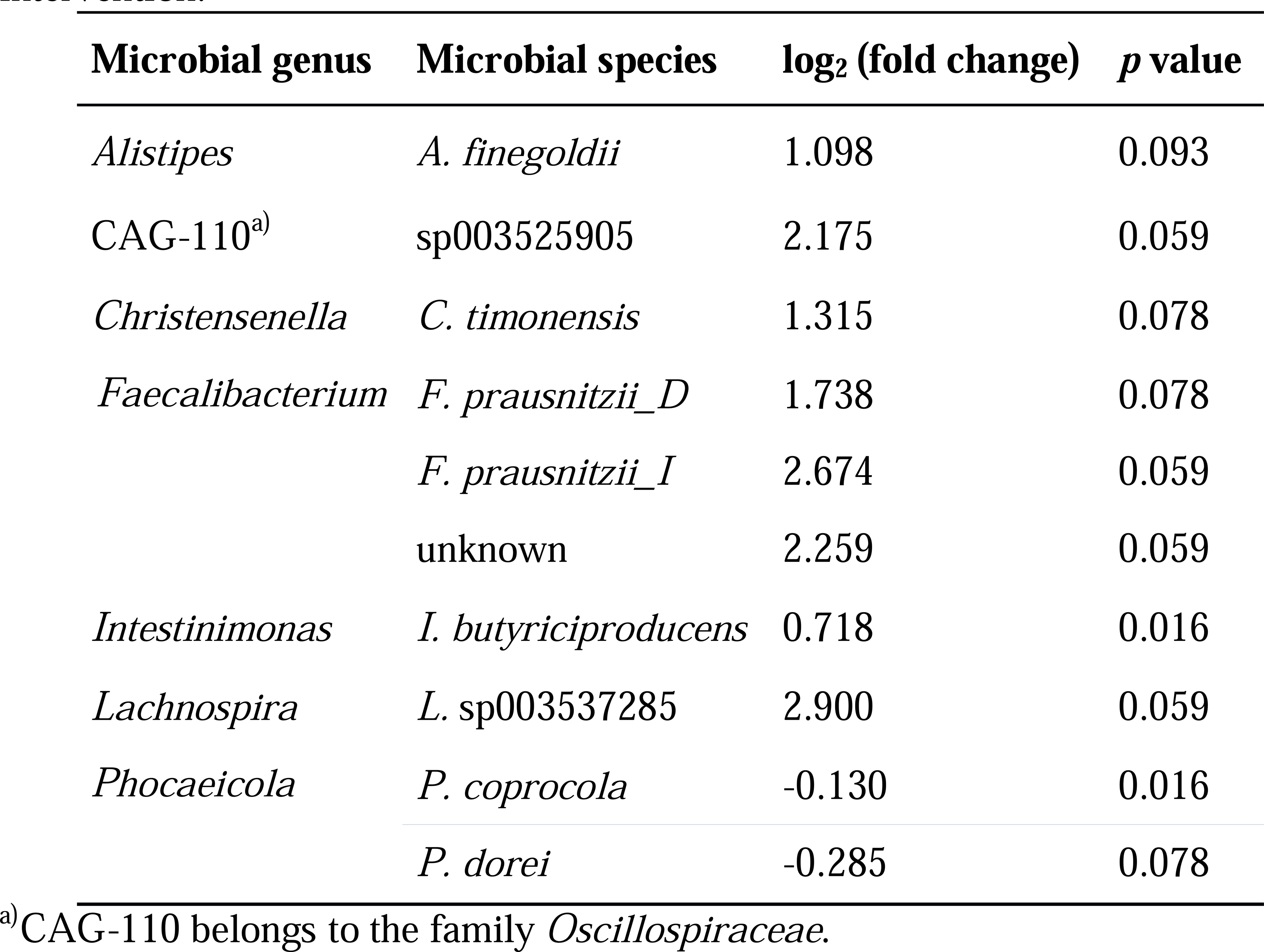
Changes in the relative abundance of microbial strains after vitamin E intervention.

**Table 2.**
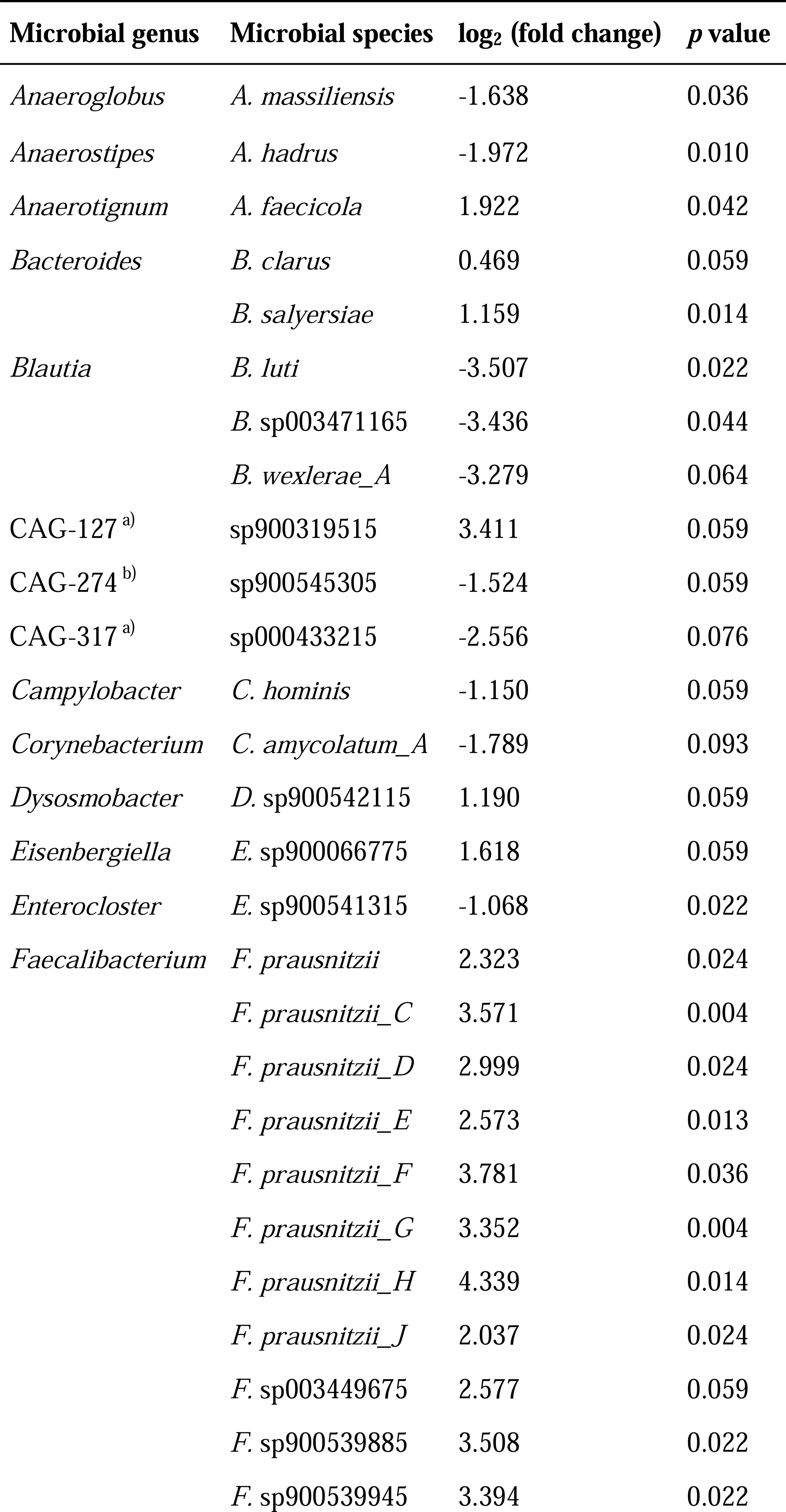
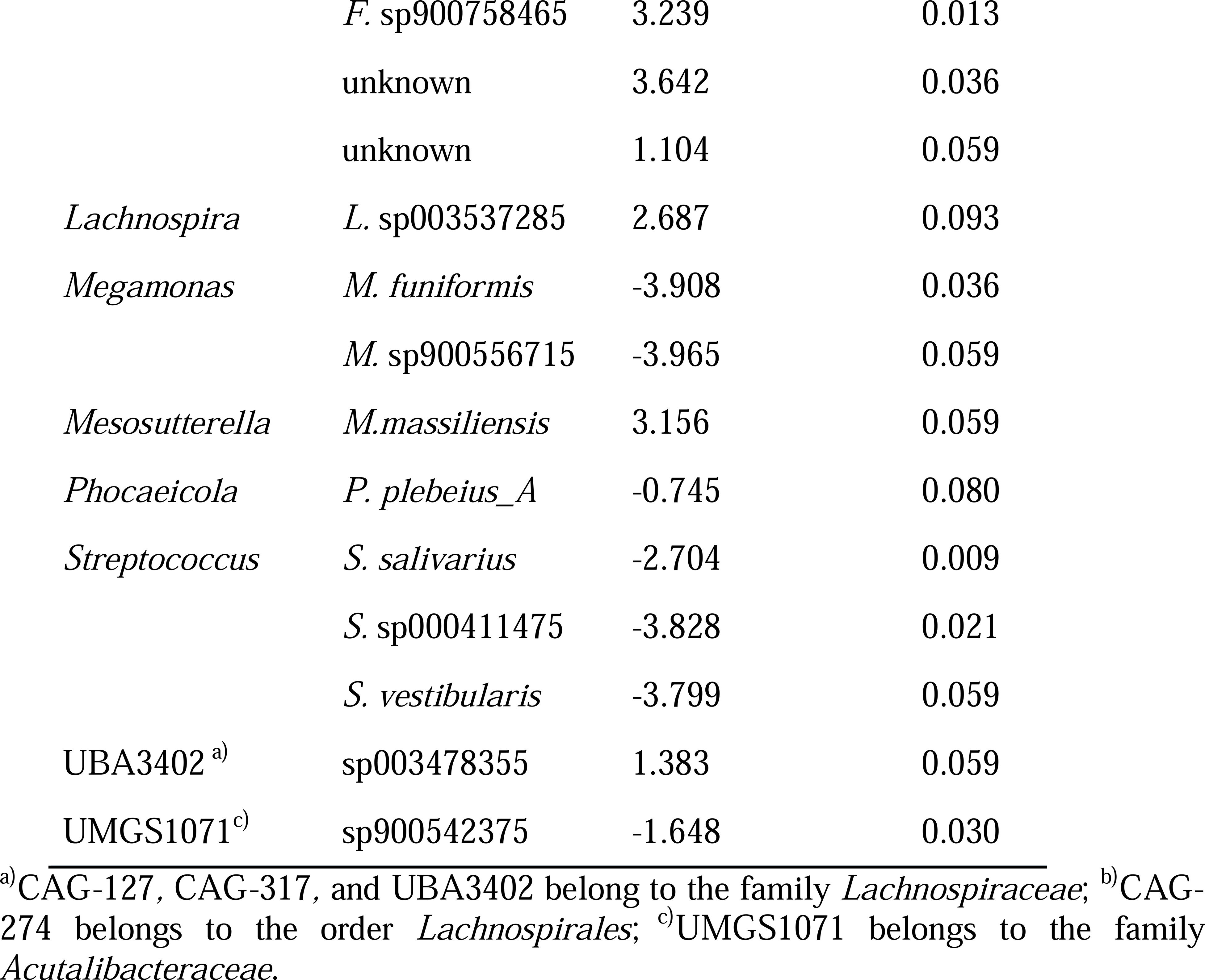
Changes in the relative abundance of microbial strains after mixed berry juice intervention.

Furthermore, we analyzed the gut microbiota composition of subjects after the withdrawal period. *Lachnospira* sp., *Faecalibacterium* spp. and *Intestinimonas butyriciproducens* maintained an upward trend during the withdrawal period (*P*_paired Wilcoxon_ < 0.1 *vs.* baseline and *P*_paired Wilcoxon_ > 0.1 *vs.* intervention; Table S6). In contrast, the relative abundance of *Streptococcus salivarius* in the juice group and *Christensenella timonensis* in the VE group was comparable to the baseline level (*P*_paired Wilcoxon_ > 0.1 *vs.* baseline and *P*_paired Wilcoxon_ > 0.1 *vs.* intervention).

We further assessed the presumed functions of the identified enriched bacterial species in the gut using KEGG functional analysis. It is known that gut microbiota can affect lipid metabolism in three main ways: biosynthesis, bioconversion and transport ^[38]^. Moreover, it is widely accepted that bile acids and SCFAs are two major microbial metabolites that affect lipid metabolism, in addition to coprostanol and exopolysaccharides. In total, 30 bacterial species were found to be increased in at least one intervention group harboring genes known to be involved in SCFA production (Figure S7A). Among the identified SCFA-producing species, 17 contained the cholelglycine hydrolase gene, which may metabolize bile acids (Figure S7B). Choloylglycine hydrolase, a bile salt hydrolase, can promote the synthesis of bile acids from cholesterol by hydrolyzing bile acid salts.^[39]^ Surprisingly, no bacterial species were found to harbor genes related to coprostanol synthesis, whereas five species could be associated with exopolysaccharide production (Figure S7C). It is noteworthy that *Lachnospira* sp003537285 was found to be increased in both intervention groups and did not have SCFAs- and exopolysaccharide-producing ability, but could hydrolyze bile salts.

### 3.4 Personalized impacts of microbiome on supplementation intervention responses

Previously, it was demonstrated that the rate of change in LDL-C levels in the VE group was related to baseline serum levels of VE. To identify microbial species that may mediate responsiveness to juice or VE supplementation and those potentially involved in the hypolipidemic effect, bacterial profiles were identified in the top and bottom three responders. The levels of LDL-C in six (66.7%) participants decreased after juice supplementation (Figure 3A). The decline rate of LDL-C levels in top responders was 22.1 ± 5.5%, whereas in bottom responders, it was −12.0 ± 8.8%. Five bacterial species were found to be differentially enriched between the top and bottom responders: *Bacteroides ovatus, Bacteroides* sp902362375*, Bifidobacterium thermophilum, Escherichia coli_D,* and *Pantoea septica* (*P*_nonparametric two-way ANOVA_ < 0.05; Table S7). The abundance of *B. thermophilum* was the highest in the top responders in both stages, whereas it was the lowest in the other groups (Figure 3B). Conversely, LDL-C levels in all seven subjects in the VE group decreased (Figure 3C). The decline rates of LDL-C levels in the top and bottom responders were 20.8 ± 10.0% and 5.0 ± 3.6%, respectively. In the VE group, the abundance of *Bacteroides xylanisolvens*, *Haemophilus parainfluenzae_L*, and *Veillonella rogosae* was the highest in the top VE responders, whereas six species had the lowest abundance, *Butyricimonas paravirosa, Bifidobacterium catenulatum, Coprococcus catus, Dorea longicatena_B, Prevotella bivia,* and CAG−317 sp000433215 (*P*_nonparametric two-way ANOVA_ < 0.05; Table S6; Figure 3D). Interestingly, *V. rogosae*, a producer of acetic acid and propionic acid ^[40]^, was only detected in the top responders, whereas its relative abundance decreased after the intervention (*P*_nonparametric two-way ANOVA_ = 0.015).

**Figure 3.**
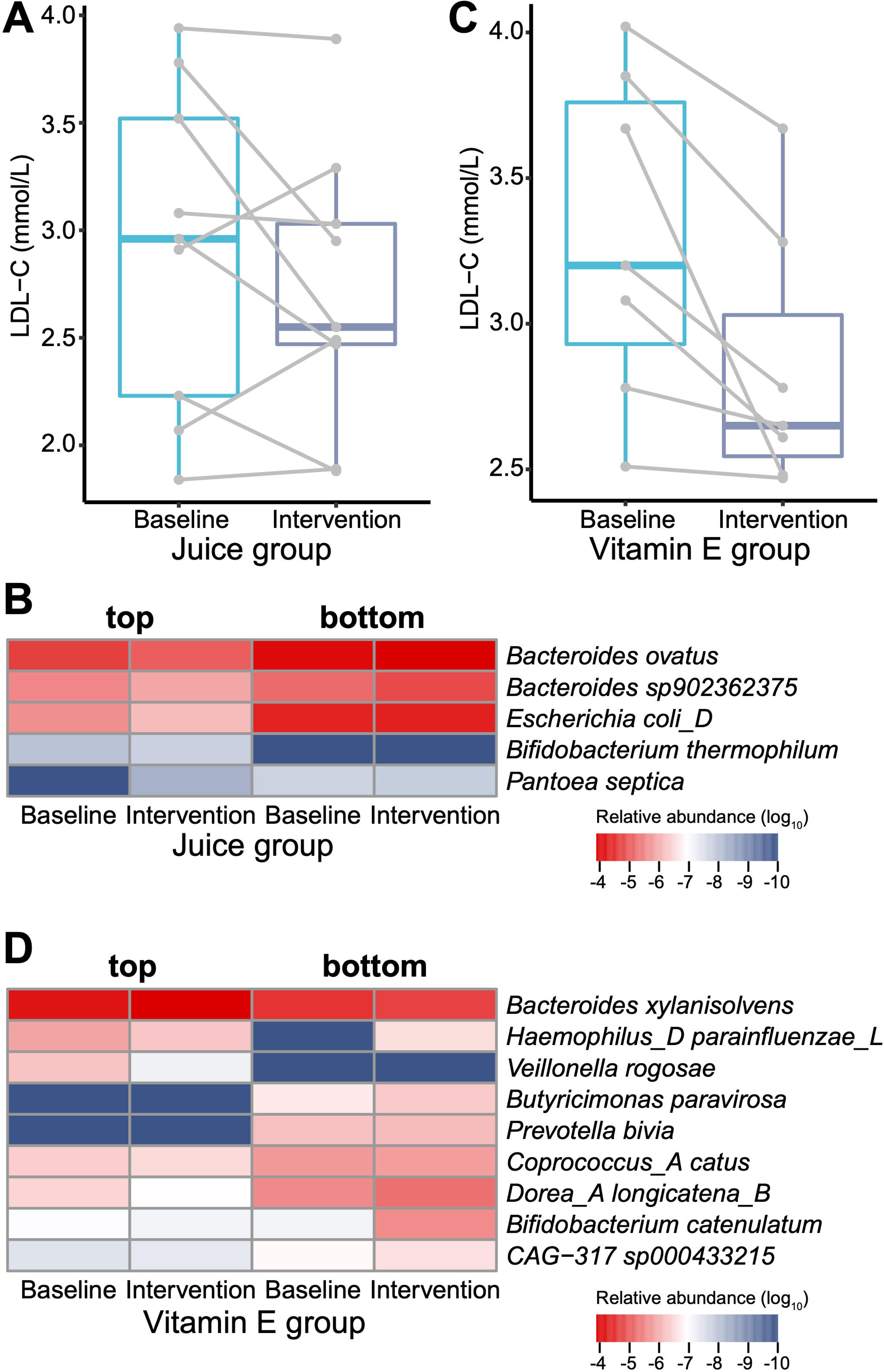
Microbial features associated with lipid-lowering effect of antioxidants. (A) Changes in low-density lipoprotein cholesterol (LDL-C) levels per individual in the mixed berry juice group. (B) Differences in the relative abundance of bacterial strains between the top and bottom three responders based on nonparametric two-way ANOVA in the mixed berry juice group. (C) Changes in LDL-C levels per individual in the vitamin E group. (D) Differences in the relative abundance of bacterial strains between the top and bottom three responders based on nonparametric two-way ANOVA in vitamin E group.

## 4. Discussion

In the present study, it was shown that the gut microbiota responded to oral supplementation with anti-aging compounds, and most bacterial groups changed after supplementation with mixed berry juice and VE, which were associated with lipid metabolism. Lower levels of LDL-C are linked to a reduced risk of cardiovascular diseases^[41]^, as well as a lower incidence of cancer^[42]^ and Alzheimer’s disease.^[43]^ A decrease in LDL-C levels in subjects receiving VE and juice supplementation has also been observed in previous studies. In a meta-analysis of randomized controlled trials (RCTs), it was found that berry consumption lowered LDL-C levels.^[44]^ Moreover, in a recent RCT with overweight women, it was proved that consumption of apple/berry juice could decrease the LDL/HDL ratio and the levels of low-density LDL (LDL3-7), rather than large LDL (LDL1, LDL2).^[45]^ Although the complete mechanism by which berries decrease LDL-C levels can be speculated that this may be due to the presence of anthocyanins derived from berries. Studies have found that anthocyanin intake can inhibit cholesteryl ester transfer protein (CEPT) and inhibit cholesterol synthesis by downregulating 3-hydroxy-3-methylglutaryl coenzyme A (HMG-CoA) reductase activation, which in turn decreases LDL-C levels.^[46]^ Conversely, VE supplementation could improve human health by increasing LDL resistance to oxidation and decreasing the cytotoxicity of oxidized LDL.^[47, 48]^ To the best of our knowledge, apart from its antioxidant capacity, the ability of VE to lower LDL-C levels has not been described. Moreover, it is worth mentioning that a previous RCT showed that LDL-C levels were not altered after long-term administration of VE in patients with dyslipidemia, which may be due to the particular gut microbiota profile of patients or the fact that patients have been taking statins.^[49]^

Moreover, it was observed that the limited alterations in the gut microbiota as a consequence of supplementation with antioxidants were not sufficient to significantly alter the gut microbiota composition in healthy individuals. Thus, the effects of juice and VE could not be attributed exclusively to antioxidant activity, as most bacterial species that changed in the two experimental groups diverged. Notably, the bacterial species related to the LDL-C-lowering effect were *Lachnospira* sp. and *Faecalibacterium* spp., which were found to be increased in both groups.^[34-37]^

*F. prausnitzii,* the sole known species within the genus *Faecalibacterium*, is one of the most abundant species in the human gut (over 5% of the total bacterial population), and is a widely recognized probiotic and the dominant SCFA producer in the gut.^[50]^ Of note, *F. prausnitzii* was found to be enriched in healthy individuals in previous studies when comparing the microbiota of patients with type 2 diabetes and lung cancer.^[9, 51]^ *Lachnospira* is a Gram-positive obligately anaerobic bacterium that ferments diverse plant polysaccharides and pectin.^[52]^ Its presence in the gut is positively associated with vegetable-rich diets (vegetarian and vegan diets), as well as with individuals taking VE. ^[53, 54]^ Of note, a decrease in the abundance of *Lachnospira* has been observed in patients with chronic kidney disease^[55]^ or immune-mediated inflammatory disease.^[56]^

Furthermore, VE supplementation increased the relative abundance of specific bacterial species in the gut of healthy individuals, such as *A. finegoldii, I. butyriciproducens,* and *C. timonensis*, but decreased the abundance of *P. coprocola* and *P. dorei* (Table 1). Previous studies have shown that *A. finegoldii* was positively associated with fecal coprostanol in humans.^[57]^ In addition, juice intervention resulted in more dramatic alterations in the gut microbiota structure; the relative abundance of certain potentially probiotic bacteria decreased (Table 2), including *B. luti*, *B. wexlerae* ^[58]^, and *S. Salivarius.*^[59]^ In both groups, as demonstrated by KEGG functional analysis, an increase in the abundance of bacterial species that harbored genes related to the production of SCFA and bile acid. Among these, *F. prausnitzii*, *I. butyriciproducens*, and *B. salyersiae* have been extensively studied, whereas other unknown species have also been identified, such as CAG-110 sp003525905, CAG-127 sp900319515 and UBA3402 sp003478355.

In addition, although the juice and VE interventions could be statistically associated with a decrease in LDL-C levels, the levels of LDL-C did not decrease in approximately one-third of the subjects in the juice group after the intervention, thus indicating a heterogeneous response to the treatment evaluated in our study. Response heterogeneity is a common phenomenon in RCT, especially in disease-related studies.^[60]^ This heterogeneity was explored in terms of baseline gut microbial profiles, and indicated that in the gut microbiota of poorly responding individuals during juice intervention, the probiotic *B. thermophilum* was depleted ^[61]^, whereas opportunistic pathogens were enriched, which included *E. coli* ^[62]^, *B. ovatus* ^[63]^ and *P. Septica.*^[64]^ It has been demonstrated that *B. ovatus* was also found to be decreased in the gut microbiota during intervention with proanthocyanidin-enriched cranberry extract.^[65]^ As for VE intervention, certain bacteria were found enriched in patients with clinical symptoms, such as *P. bivia* in the vaginal tract of infected patients ^[66]^, *C. catus* in type 2 diabetic patients ^[67]^ and *D. longicatena* in obese patients ^[36]^, which were less enriched in high responding subjects. Beneficial bacteria were observed in individuals with top or bottom responses (Figure 3D). Thus, it can be speculated that the enrichment of disease-associated bacteria in the gut, rather than that of probiotics, was the key to the lipid-lowering effect.

Previous studies have shown that the effect of an intervention with dietary supplementation is time-dependent and generally subsides after one to two times the corresponding amount of time the intervention lasted.^[68, 69]^ In the present study, alterations in the abundance of *Lachnospira* sp., *Faecalibacterium* spp. and *I. butyriciproducens* lasted for over three months after the intervention had ceased. In this context, two scenarios can be drawn: firstly, the three-month intervention time was sufficient to enable these bacterial species to become dominant or be eliminated in competition with new dominant bacterial species in the gut; secondly, these fluctuations may disappear completely after a long post-intervention time. Future research is needed to elucidate these aspects and confirm our hypotheses.

Herein, a three-month intervention in Chinese individuals was conducted involving supplementation based on three types of antioxidants, and information during the post-intervention period was also obtained. We sought to shed light on the following aspects of supplementation: i) whether long-term supplementation could provide beneficial effects without the need to implement changes in the diet; and ii) the duration of the effects resulting from the intervention. However, certain limitations were observed: i) although the three-month intervention time may have enabled the elucidation of the effects of the supplementation schemes throughout a wide time frame, the three-month intervention period may have contributed to the increase in withdrawal rate, since only 60 percent of participants who were initially enrolled in the study completed the trial. Therefore, to avoid the occurrence of false-negative results caused by a lower number of fecal samples than initially expected, we did not perform FDR correction on *p*-values ^[70]^ in gut microbiota analysis. ii) *in vivo* experiments in mice or with artificial gastrointestinal tract models were not included, which could have provided more information about the underlying mechanisms of juice or VE.

Collectively, the findings of the present study revealed that mixed berry juice, VE, and GSE may not act as anti-aging compounds, as expected, but juice and VE could benefit cardiovascular function by lowering LDL-C levels. Moreover, it can be proposed that their effects are indirectly mediated through their impact on the human gut microbiota. Finally, two SCFA-producing bacteria, *Lachnospira* sp. and *Faecalibacterium* spp., may play important roles in the gut microbiota and deserve further study.

## Author Contributions

B.W.C. and Y.X.L. contributed equally. B.W.C. and T.L. designed the study. L.X. and T.L. supervised the study. H.F.Z., H.Y.C. and X.J.H. collected the data. B.W.C. performed the bioinformatic/statistical analyses with the assistance of Z.M.L. B.W.C. and Y.X.L. wrote the first draft of manuscript. Z.M.L. and T.L. reviewed the manuscript. B.W.C., L.X. and T.L. made the final decision. T.L. acquired the grant funds. Y.H., C.N., and X.J. provided resource. All authors had full access to the final version of the manuscript and agreed to its submission for publication.

## Data Availability

The data that support the findings of this study are available from the CNGB Sequence Archive (CNSA) of the China National GeneBank Database (CNGBdb). Restrictions apply to the availability of these data, which were used under license for this study. Data are available from the corresponding authors with the permission of CNGBdb.

## Abbreviation list

BWA: burrows-wheeler aligner
CNSA: CNGB sequence archive
CNGBdb: china national genebank database
CEPT: cholesteryl ester transfer protein
FDR: false discovery rate
GSE: grape seed extract
HMG-CoA: 3-hydroxy-3-methylglutaryl coenzyme A
KEGG: kyoto encyclopedia for genes and genomes
KO: KEGG orthology
LDL-C: low-density lipoprotein cholesterol
LDL3-7: low-density LDL
LDL1, LDL2: large LDL
NLR: neutrophil-to-lymphocyte ratio
PERMANOVA: permutational multivariate analysis of variance
PCoA: principal coordinates analysis
RCTs: randomized controlled trials
SCFAs: short-chain fatty acids
SII: systemic immune-inflammation index
UHGG: unified human gastrointestinal genome
VE: vitamin E

## Acknowledgments

We would like to thank our colleagues of BGI-Research and the volunteers who participated in the study for their dedication, commitment, and contribution. This study was supported by the National Key Research and Development Program of China (2020YFC2008002), Guangdong Province International, Hong Kong, Macao, and Taiwan High-end Talent Exchange Project “Senior Talents” (2021A1313030024).

## Supplementary materials

Supplemental figures and tables are provided as supplemental materials.

