## Supplemental Figure 1-7 for "Supplementation with Berry Juice and Vitamin E Ameliorates Blood Cholesterol Level and Alters Gut Microbiota Composition"

**Supplementary Figures**

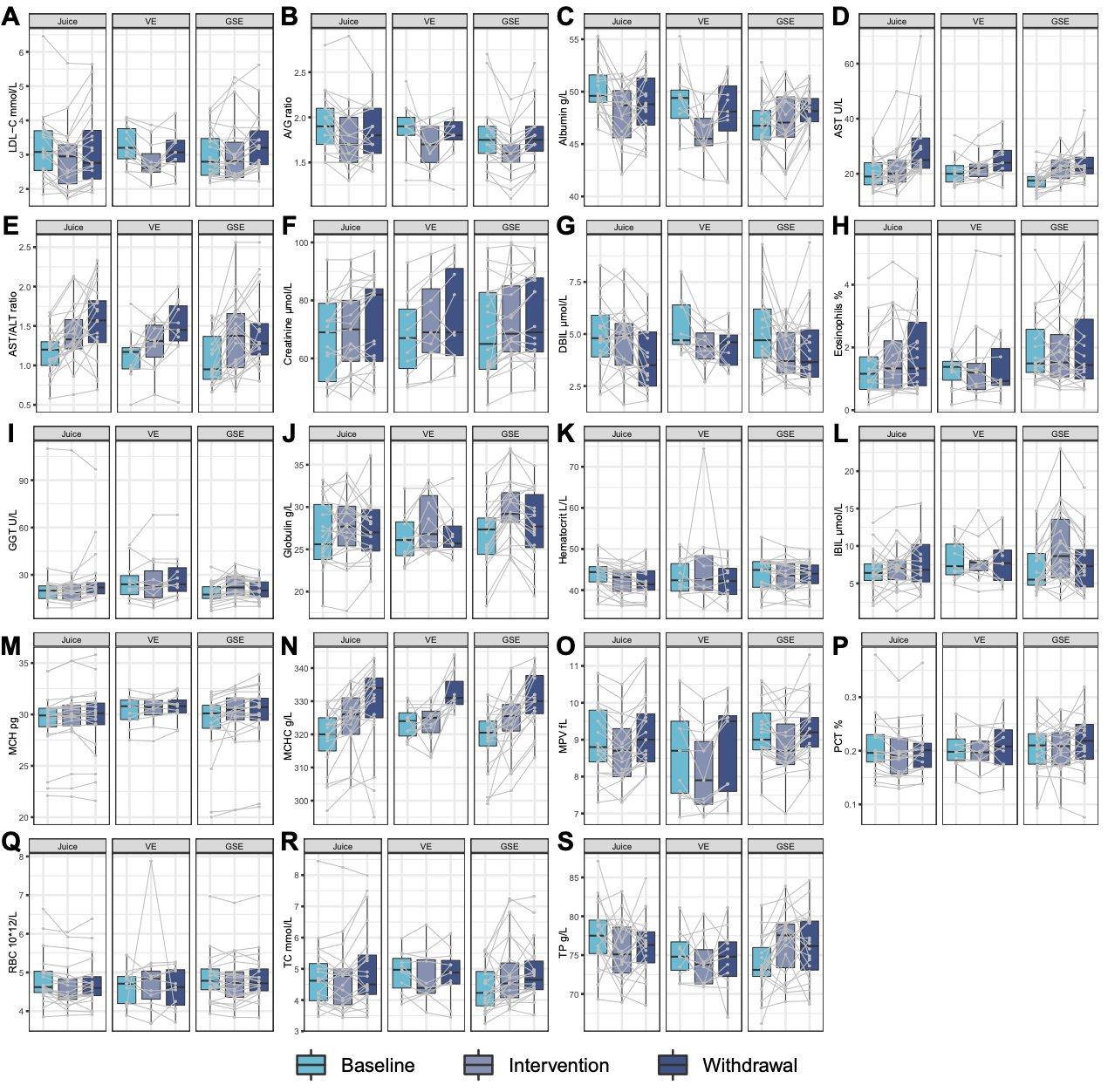

**FIG S1** Changes in blood test levels. (A) low-density lipoprotein cholesterol (LDL-C); (B) albumin/globulin (A/G) ratio; (C) Albumin; (D) Aspartate transferase (AST); (E) AST/alanine transaminase (ALT) ratio; (F) Creatinine, (G) Direct bilirubin (DBIL); (H) Eosinophils; (I) Gamma-glutamyl transferase (GGT); (J) Globulin; (K) Hematocrit; (L) Indirect bilirubin (IBIL); (M) Mean cell hemoglobin (MCH); (N) Mean corpuscular hemoglobin concentration (MCHC); (O) Mean platelet volume (MPV); (P) Plateletcrit (PCT); (Q) Red blood cell (RBC); (R) Total cholesterol (TC); (S) Total protein (TP). Juice, mixed berry juice; VE, vitamin E; GSE, grape seed extract.

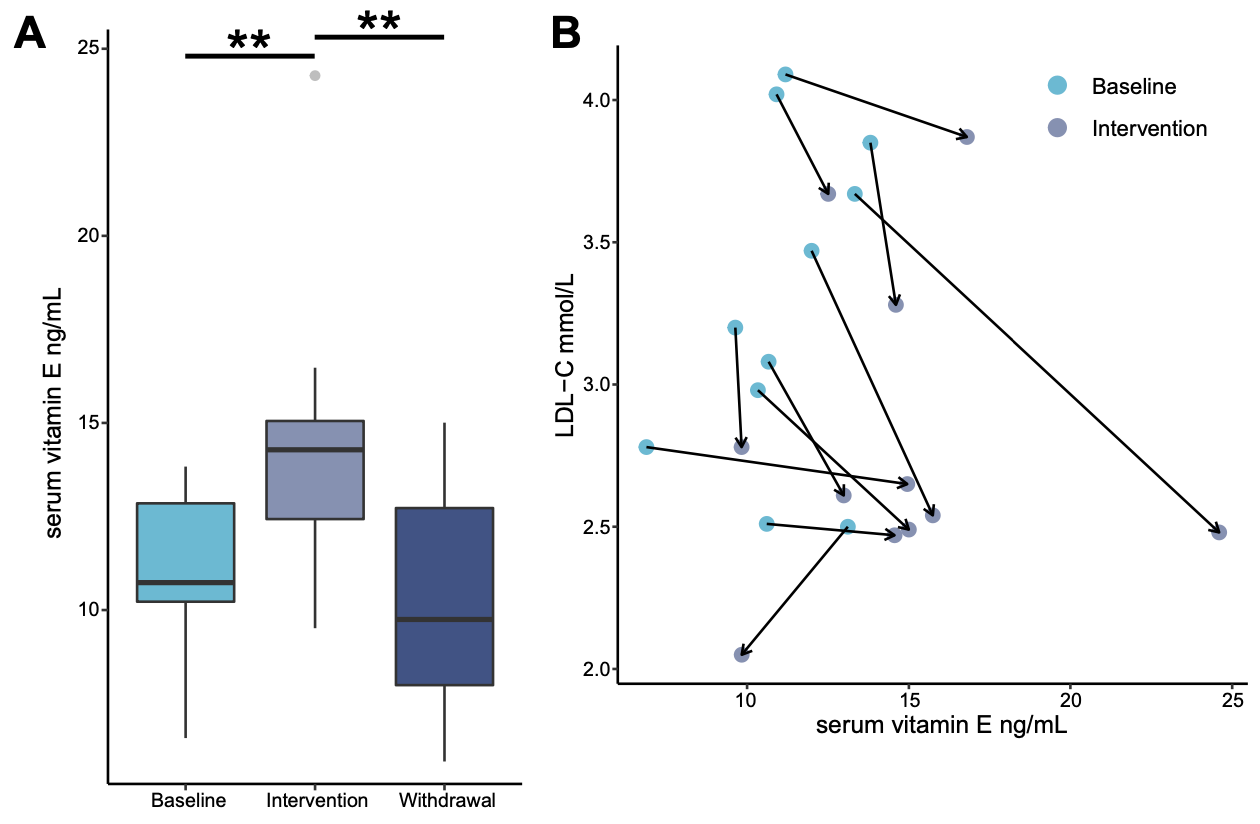

**FIG S2** Changes in serum vitamin E levels in healthy individuals receiving vitamin E supplementation. (A) Serum vitamin E levels at different intervention stages. Paired Wilcoxon rank-sum test, ***P* < 0.01. (B) Changes in serum vitamin E levels and low-density lipoprotein cholesterol (LDL-C) levels before and after vitamin E intervention.

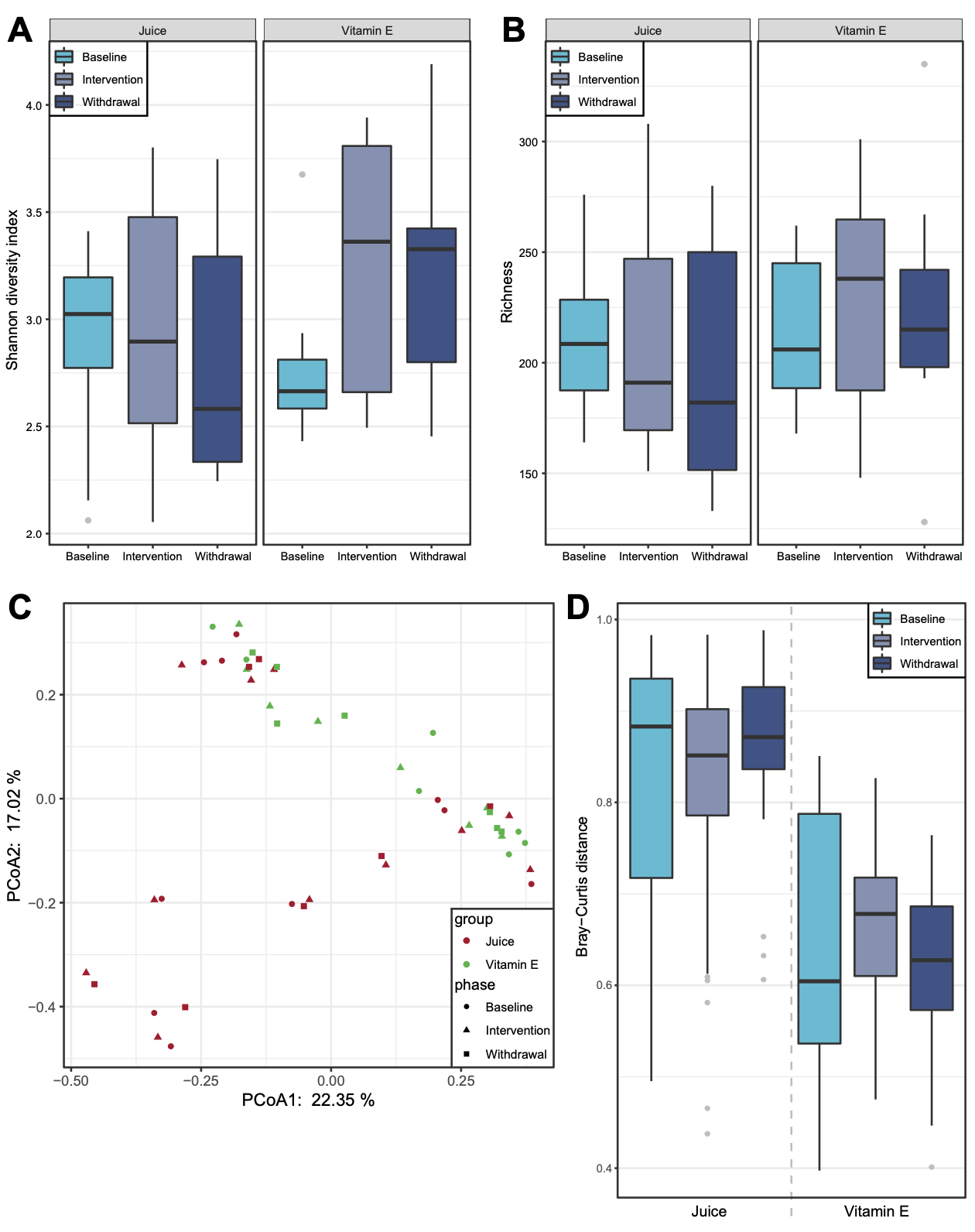

**FIG S3** Composition and diversity of gut microbiota in healthy individuals receiving supplementation with mixed berry juice group and vitamin E. (A-B) Distribution of Shannon diversity index and richness. (C) Principal coordinates analysis (PCoA) of gut bacterial species based on Bray−Curtis distance. (D) Distribution of Bray−Curtis distance.

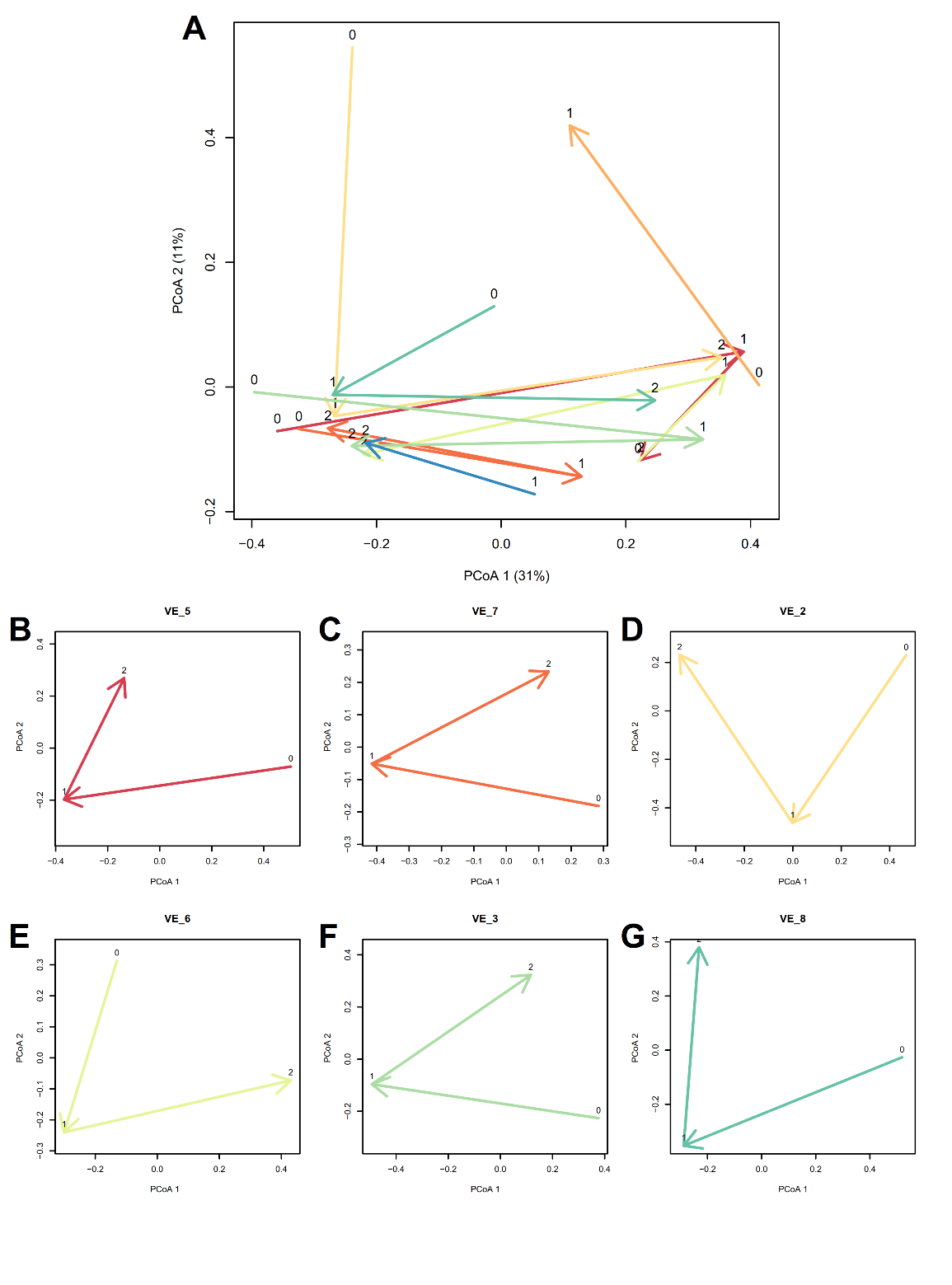

**FIG S4** Trajectory analysis of changing trends in the gut microbiota during the withdrawal period in healthy individuals receiving supplementation with vitamin E. (A) Analysis of all surveyed individuals. (B-G) Changes in each participant in the study whose data were collected at three different sampling points. PCoA, principal coordinates analysis; VE, vitamin E.

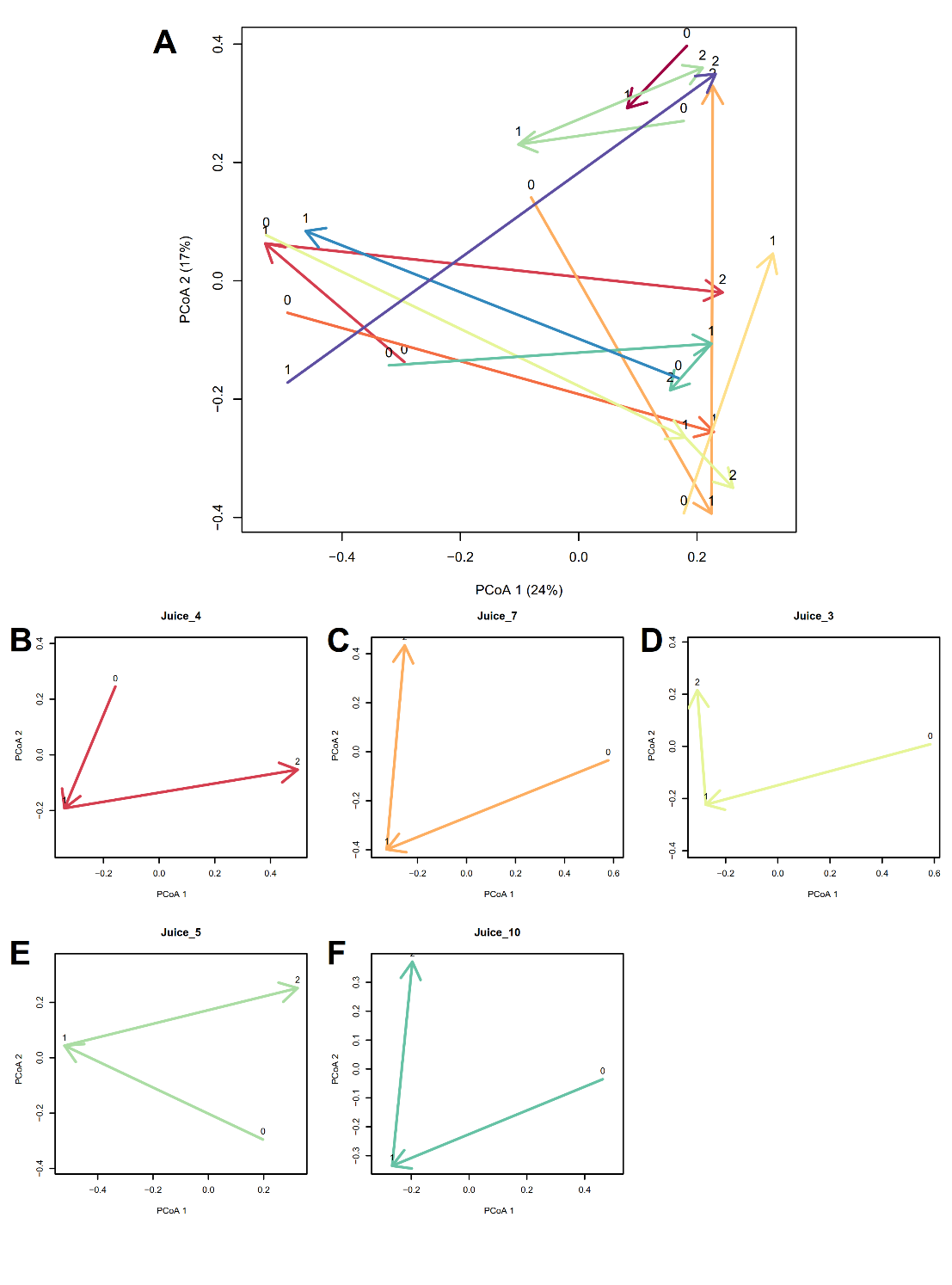

**FIG S5** Trajectory analysis of changing trends in the gut microbiota during the withdrawal period in healthy individuals receiving supplementation with mixed berry juice. (A) Analysis of all surveyed individuals. (B-G) Changes in each participant in the study whose data were collected at three different sampling points. PCoA, principal coordinates analysis; VE, vitamin E.

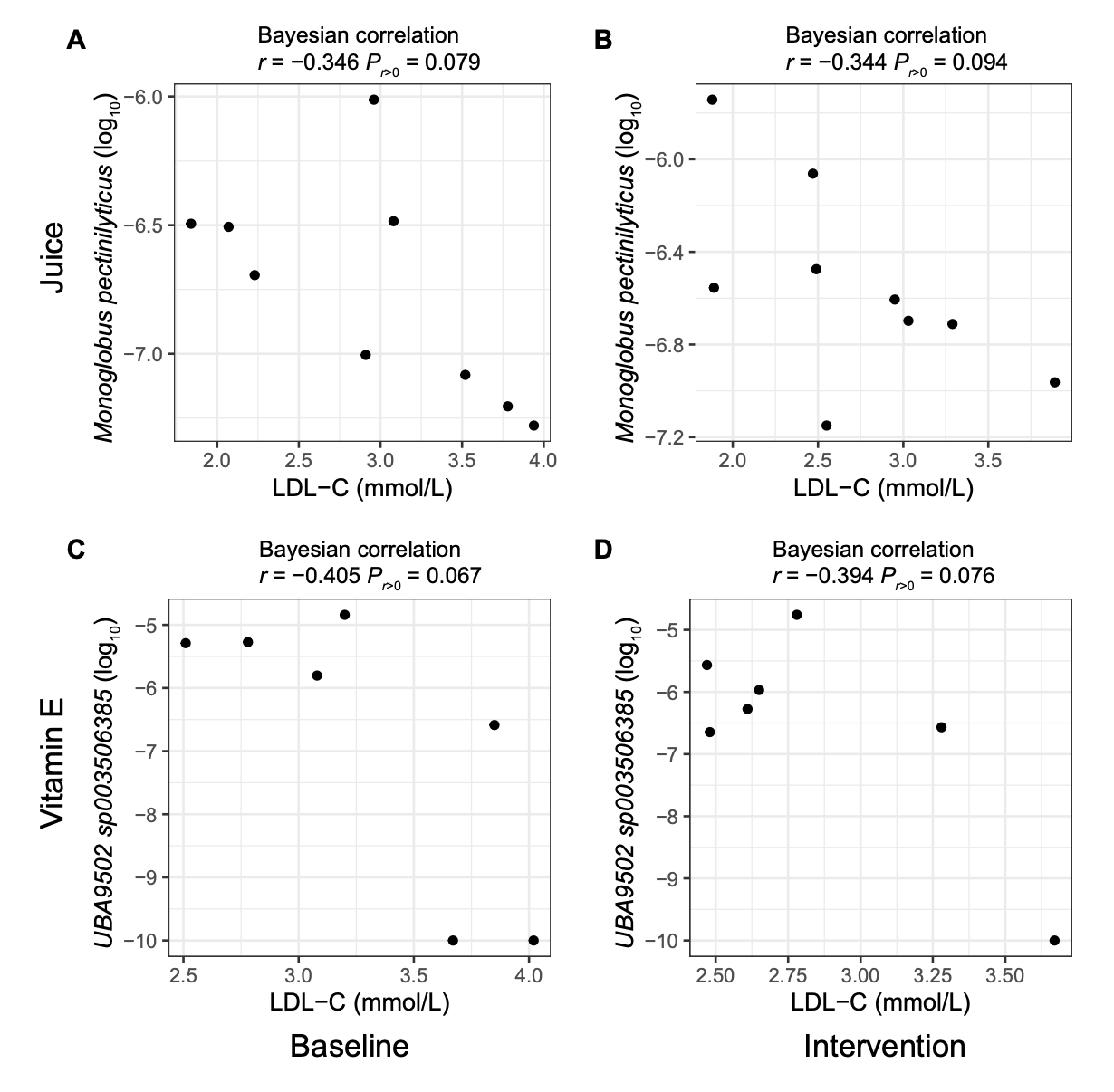

**FIG S6** Bayesian correlation analysis between the relative abundance of bacterial strains and levels of low-density lipoprotein cholesterol (LDL-C) among individuals from mixed berry juice or vitamin E intervention groups.

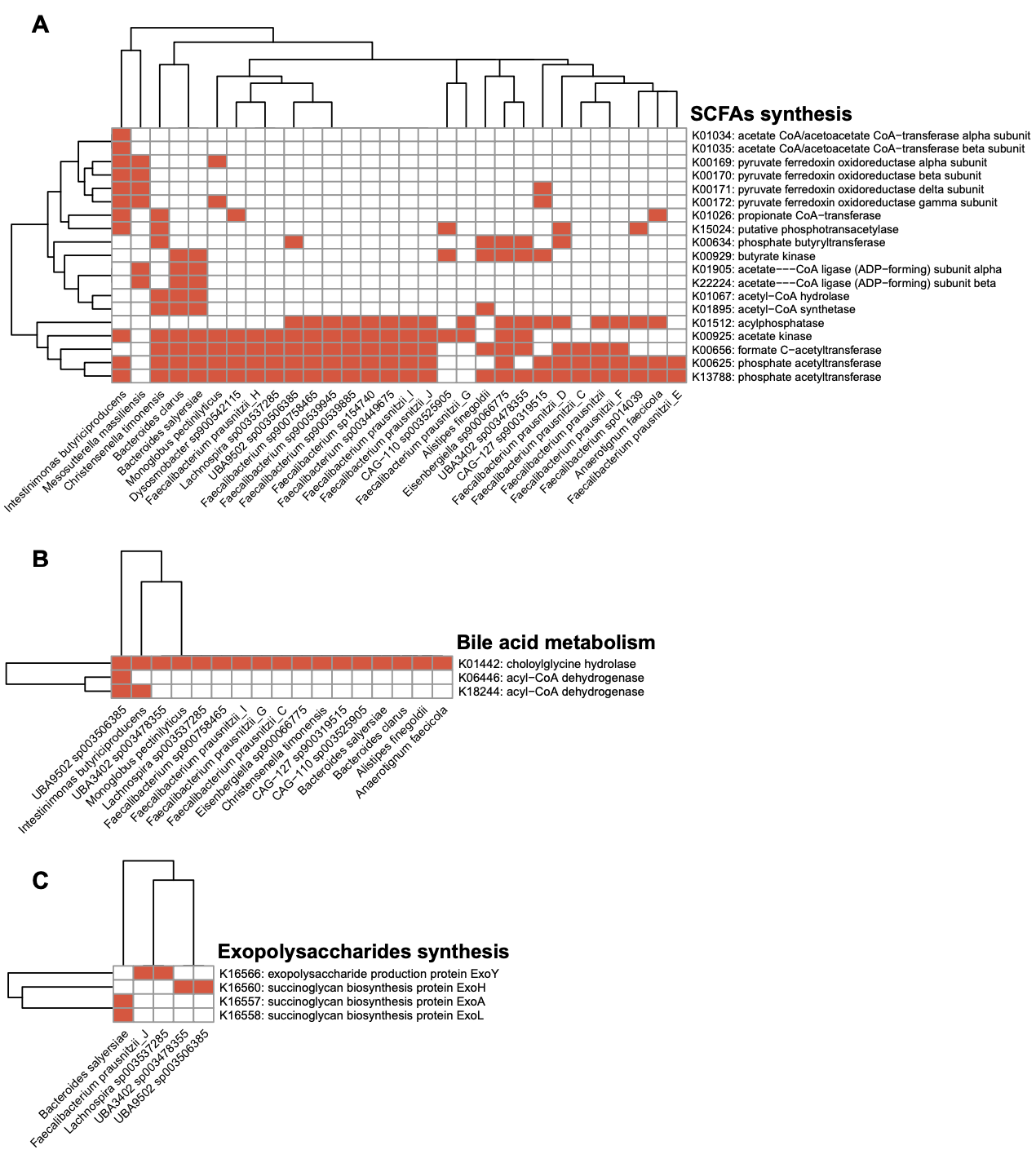

**FIG S7** KEGG functional analysis of bacterial genes involved in cholesterol metabolism found in representatives of the gut microbiome of individuals receiving supplementation with vitamin E or mixed berry juice. (A) Genes annotated within short chain fatty acids (SCFAs) biosynthetic pathways. (B) Genes annotated within bile acids metabolic pathways. (C) Genes annotated within exopolysaccharides biosynthetic pathways.
